# Associations between Demographic Characteristics, Perceived Threat, Perceived Stress, Coping Responses and Adherence to COVID-19 Prevention Measures among Healthcare Students in China: A Cross-Sectional Survey with Implications for the Control of COVID-19

**DOI:** 10.1101/2020.07.15.20154997

**Authors:** Anson Chui Yan Tang, Enid Wai Yung Kwong, Liangying Chen, Winnie Lai Sheung Cheng

**Author notes:** Correspondence to: Anson CY Tang, 16/F Tung Wah College, 31 Wylie Road, The Hong Kong SAR, China.

## Abstract

**Objectives:** To investigate the associations between demographic characteristics, perceived threat, perceived stress, coping responses and adherence to COVID-19 prevention measures in Chinese Healthcare students.

**Design:** A cross-sectional survey collecting data in Hong Kong and Fujian Province of China. Self-administered questionnaires were collected via online platform in April 2020.

**Participants:** A convenience and snowball sample of 2706 students aged 18 years or older and studying a healthcare programme in Hong Kong or Fujian.

**Setting:** Students were recruited in tertiary education institutions/universities in Hong Kong and Putian (a prefecture-level city in eastern Fujian province). The institutions offered various healthcare programmes in degree or sub-degree levels.

**Main outcome measures:** Compliances to social distancing and personal hygiene measures were assessed by 10-item Social Distancing Scale and 5-item Personal Hygiene Scale respectively. Path analysis was performed to identify factors associated with the compliance outcomes.

**Results:** The participants reported high compliances to both social distancing and personal hygiene measures. Confidence to manage the current situation, wishful thinking and empathetic responding directly predicted compliance to social distancing (*β*=-0.31, p<0.001; *β*=0.35, p=0.015; *β*=0.33, p<0.001 respectively) and personal hygiene measures (*β*==-0.16, p<0.001; *β*=0.21, p<0.001; *β*=0.16, p<0.001 respectively). Gender, geographical location, and clinical experience were the only three demographic variables having direct and/or indirect effects on social distancing and personal hygiene measures. The final model constructed demonstrated a very good fit to the data (Chi-square *X*^2^=27.27, df=17, P=0.044; *X*^2^/df=1.61; GFI=0.998, CFI=0.997, TLI=0.992, RMSEA=0.015).

**Conclusions:** The predictive model constructed in this study is the first one to explore factors associating with the compliance to infection control measures in healthcare students amid the COVID-19 outbreak. The findings suggest that students who are male, habituate in Hong Kong, have more clinical experience and weak confidence to manage the threat tend to have lower compliance to social distancing and personal hygiene measures. Wishful thinking, contrasting to previous studies, was first found to positively associate with adherence to COVID-19 control measures.

## Introduction

Since China first identified and reported a pneumonia case in Wuhan to the World Health Organization (WHO) Country Office on 31 December 2019, COVID-19 has infected over 10 million people in more than 46 countries, resulting in 499,913 deaths as of 30 June 2020.^1^ The extent of community spread of COVID-19 is significantly greater than that for previous viral outbreaks such as severe acute respiratory syndrome (SARS) in 2003, H1N1 influenza in 2009, and Middle East respiratory syndrome (MERS) in 2012 and 2015. The highly contagious nature of COVID-19, even during the incubation period, favours the insidious and rapid transmission of SARS-CoV-2 (the virus responsible for the COVID-19 outbreak) from person to person via respiratory droplets and contact, as asymptomatic people are unaware of being infected.^2-4^ In efforts to swiftly control the continuous spread of the virus, many countries have been launching a series of stringent infection prevention and control measures (ICPs), such as mandatory quarantine, massive lockdowns, border control, and suspending schooling to seal off transmission. However, these aggressive ICPs have unprecedentedly restricted daily living and hampered global economic activities. Reports warn that the COVID-19 pandemic will shrink global GDP by almost one percent in 2020,^5^ hitting people’s livelihoods hard due to the resulting depression of the employment market.^6,7^ To mitigate the socioeconomic impact of the pandemic, governments have been gradually resuming normal social and economic activities along with less restrictive but essential ICPs to control the spread of the virus.^8^

### Rationale of the proposed study

At present, implementing essential ICPs is the sole approach to mitigate virus spread in the community. WHO and national health agencies have compiled a recommended list of COVID-19 ICPs for the general public to follow, including social distancing, avoiding crowds, wearing face coverings, handwashing with soap and water or using hand sanitizer with at least 60% alcohol, and regular cleaning and disinfection of touched surfaces.^3,4,9^ Reports related to numerous community cluster infections have indicated the importance of strict compliance with the recommended ICPs to minimise viral transmission. For instance, the nightclub cluster in South Korea, which was initiated by a man visiting several nightclubs without wearing a mask, led to a resurgence of new infections in South Korea.^8^ Understanding the underlying factors that determine people’s compliance with ICPs in a community setting is therefore a pressing need.^10^ Evidence regarding compliance with ICPs in the context of the COVID-19 pandemic is currently underreported. With reference to previous studies concerning factors related to ICP adherence during the earlier SARS and H1N1 outbreaks, notable regional and demographic differences were observed in the adherence to the ICPs by the general public.^10-15^ Older, more educated females are generally reported as having higher compliance with the ICPs.^10,12-14^ Moreover, higher ICP compliance is also associated with those who have a higher anxiety level, higher perceived risk of infection, better knowledge about the infection, and belief in the effectiveness of the recommended ICPs.^10,12-15^ Greater trust in authorities has also been found to favour engaging in ICPs.^25^ This study targeted healthcare students, a group that is currently under-investigated regarding their compliance with ICPs. Relevant studies have often targeted healthcare workers during virus outbreaks.^16-20^ According to the scanty evidence available, healthcare students were not as knowledgeable and competent as their seniors regarding ICPs. Even medical students’ significant improvement in hand hygiene practice after experiencing SARS was still considered insufficient.^21^ In contrast, nursing students were found to exercise moderate compliance with universal precautionary measures when they possessed sufficient knowledge about infection control precautions.^22^ When healthcare students participate in internship/practicum in hospitals/clinics, insufficient compliance with ICPs on their part may facilitate viral spread from community to hospital, or vice versa. Therefore, the present study aims to investigate the determining factors associated with compliance with COVID-19 ICPs by healthcare students to fill in gaps in the existing body of knowledge and help stakeholders devise effective public health interventions to enhance healthcare students’ compliance with ICPs.

### Hypothesised model

For the purpose of understanding adherence to COVID-19 ICPs, a hypothesised health behaviour model was formulated based on the health belief model^23^ and transactional model of stress and coping^24^ to explain the associations among potential contributing factors and compliance with ICPs during the COVID-19 pandemic (Figure 1). Engaging in behaviours that promote health (i.e. COVID-19 ICP behaviour) is influenced by the perceived threat of COVID-19, such as the chance of becoming infected, the prognosis, and the perceived stress level, which is affected by the degree of perceived threat. An increase in the stress level will bring about an evaluation of coping responses to overcome the stress, which will eventually provoke a series of health behaviours to cope with stress. For the general public, specifically, WHO and national health agencies have recommended social distancing and personal hygiene measures to prevent COVID-19 transmission in the community. Examples of social distancing are avoiding crowded places and refraining from shaking hands, while measures related to personal hygiene measures include wearing a mask and handwashing with hand sanitizer. For the coping response in this study, we chose wishful thinking and empathetic responding, which were found to be commonly adopted by individuals during outbreaks of SARS and West Nile virus.^25,26^ Wishful thinking is a type of emotion-focused coping response that refers to an individual’s effort to cognitively escape from or avoid a situation by simply fantasising or hoping the situation will go away or will somehow be over. Evidence has shown that wishful thinking is often associated with negative health outcomes such as poor adjustment to illness.^27^ Empathetic responding is a relationship-focused coping response in which the individual attempts to understand what others are experiencing and makes an effort to offer support and assistance. Lee-Baggley et al.^25^ found wishful thinking was positively associated with avoidant health behaviours such as eschewing public areas and people during a SARS outbreak. Conversely, empathetic responding was associated with proactive prevention behaviours such as mask wearing and exerting caution in personal hygiene.^25,26^ The previous literature has suggested that demographic characteristics play a significant role in people’s health behaviours. Consequently, demographic characteristics were included in the hypothesised model as they were believed to indirectly influence engagement in ICPs through perceived threat, perceived stress, and coping responses. Path analysis was adopted to verify the hypothesised associations among the studied variables in this model.

**Figure 1.**
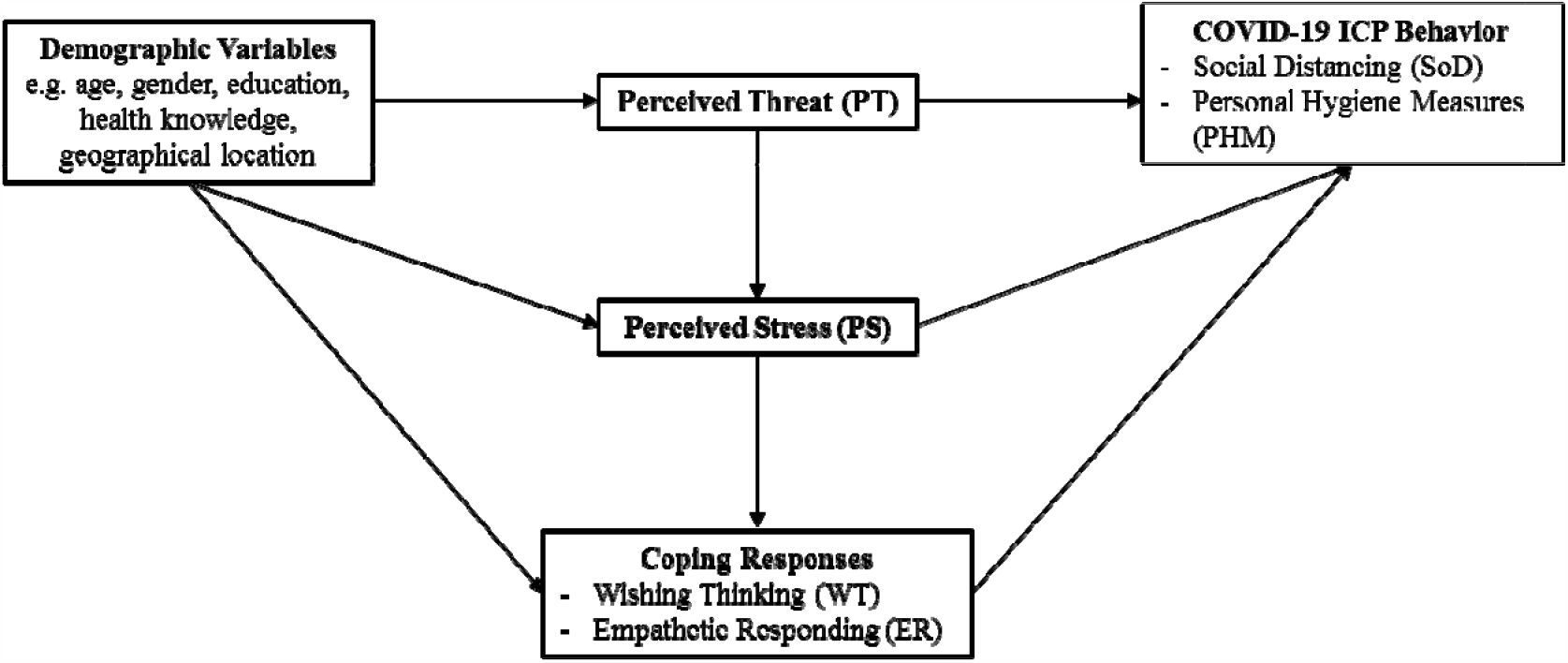
Hypothesized model of the associations among the study variables.

### Objective

The objective of this study was to investigate the associations between demographic characteristics, perceived threat, perceived stress, coping responses, and adherence to COVID-19 ICPs in Chinese healthcare students using path analysis. The main hypothesis was that demographic characteristics, perceived threat, perceived stress, and coping responses associate with social distancing and personal hygiene measures.

## Methods

### Study design and setting

We conducted a cross-sectional study, collecting data in Hong Kong and Fujian in China. In Hong Kong, the samples included students from two universities and one tertiary education institution. Students from three tertiary education institutions in Putian, a prefecture-level city in eastern Fujian Province, were also a part of the study. These universities/tertiary education institutions offer various healthcare programmes at degree or sub-degree levels such as nursing, Chinese medicine, physiotherapy, and occupational therapy. Students undertaking these programmes must take taught courses and undergo clinical practicums in hospital and community settings. Practicum hours vary among programmes. Eligible students were recruited via online platforms in April 2020. Participants completed self-administered online questionnaires that collected data on demographic characteristics, perceived threat, perceived stress level, use of wishful thinking, and empathetic responding and adherence to social distancing and personal hygiene measures in response to COVID-19. Completing the entire questionnaire required about 15 minutes. The participants were informed of the objectives and procedures of the study and their rights of participation before beginning the survey. Returning the completed questionnaire implied consent to participate in the study. The manuscript was written according to the STROBE guideline for observational studies.^28^

### Participants

Eligibility criteria included fulltime students aged 18 years or older and engaged in one of the healthcare programmes offered in the universities or the tertiary education institutions for this study. Such programmes included general/obstetric nursing, medical laboratory science, Chinese medicine, radiation therapy, pharmacy, occupational therapy, and physiotherapy. Students were ineligible if they were studying in a programme other than those listed. Mass emails were sent to the students via school emails to invite them and their friends to complete the online questionnaire. Total participants numbered 2,706, and the number of parameters to be estimated in the predictive model was 38. This sample size was adequate for modelling, in which the ratio of sample size to estimated parameters should be 5:1.^29^

### Outcome measures

#### Compliance with social distancing and personal hygiene measures

Our main outcome of interest was adherence to COVID-19 ICPs, comprising social distancing (SoD) and personal hygiene measures (PHM). The scales for measuring SoD and PHM were developed based on the WHO’s and Centers for Disease Control and Prevention’s guidelines for preventing and controlling the spread of COVID-19 for the public and individuals,^3,4^ along with questionnaires developed by the SARS Collaborative Research Group in 2004.^25^ SoD was measured using ten items, including “avoided travel to COVID-19 infected areas,” “avoided eating in restaurants,” and “avoided shaking hands.” PHM was measured by five items, such as “wearing a mask,” “washing hands more often,” and “using disinfectants.” Participants were directed to rate their likelihood of performing the actions described in the items to prevent COVID-19 on a 5-point Likert scale, where 1 was “Very unlikely,” and 5 was “Very likely.” The total SoD score and PHM score were the aggregation of the corresponding item scores. The higher the score was, the more likely the respondent was to engage in the corresponding measures.

#### Perceived threat

Perceived threat (PT) was measured by four items extracted from the perceived threat scale originally used to measure an individual’s perceived threat due to SARS.^26^ The participants were asked to rate the extent to which four statements were true for them at the current moment on a 4-point Likert scale, ranging from “Not at all” (1) to “A great extent” (4). Item examples are “I feel nervous about getting COVID-19,” and “COVID-19 is threatening my health.” A summation of the items’ scores formed the total PT score. The higher the score, the greater was the perceived threat to the individual.

#### Perceived stress

Perceived stress (PS) was measured by the perceived stress scale (PSS-10) developed by Cohen and Williamson.^30^ The PSS-10 is a 10-item self-reported questionnaire that asks respondents to rate the frequency of their feelings and thoughts about life events and situations over the previous month using a 5-point scale ranging from 0 (“Never”) to 4 (“Very often”). According to our results after confirmatory factor analysis (Table 1), the PSS-10 was divided into two subscales that measured stress level (PS1) and confidence about managing the current situation (PS2). PS1 was measured by six items, such as “In the last month, how often have you been upset because of something that happened unexpectedly?” and “In the last month, how often have you found that you could not cope with all the things that you had to do?” A higher score indicated greater stress. PS2 was measured by four items, including “In the last month, how often have you felt confident about your ability to handle your personal problems?” and “In the last month, how often have you felt that things were going your way?” PS2 was inversely scored, meaning that a higher total PS2 score indicated lower confidence about handling the current situation.

**Table 1.** Psychometric analyses of the scales used in the present study

| Scales | Variables | Item numbers<br>on variable | Factor<br>loadings | Cronbach's alpha in<br>the testing sample | Possible<br>range |
| --- | --- | --- | --- | --- | --- |
| Social Distancing<br>Scale | Social Distancing (SoD) | 10 | 0.58-0.84 | 0.93 | 5-50 |
| Personal Hygiene<br>Scale | Personal Hygiene Measures<br>(PHM) | 5 | 0.50-0.96 | 0.83 | 5-25 |
| Wishing thinking<br>Scale | Wishing Thinking (WT) | 2 | 0.73-0.77 | 0.70 | 2-8 |
| Empathetic<br>Responding Scale | Empathetic Responding (ER) | 6 | 0.61-0.92 | 0.92 | 6-24 |
| Perceived Stress<br>Scale | Perceived Stress (PS) | Domain 1 (PS1):<br>6 | 0.58-0.89 | 0.90 | 0-24 |
|  |  | Domain 2 (PS2):<br>4 | 0.62-0.85 | 0.79 | 0-16 |
| Perceived Threat<br>Scale | Perceived Threat (PT) | 4 | 0.51-0.72 | 0.70 | 4-16 |

#### Coping responses

Wishful thinking (WT) and empathetic responding (ER) were measured by two items taken from the wishful thinking subscale of the ways of coping questionnaire^31^ and six items taken from the relationship-focused coping scale,^32^ respectively. Participants rated the items from the two scales on a 4-point Likert scale, ranging from “Not at all” (1) to “A great deal” (4) to indicate the extent to which they had managed their concerns or fears about COVID-19 through wishing or helping others. The two items for measuring WT were “Wishing COVID-19 would go away or somehow be over with” and “Hoped a miracle would happen.” Examples of items measuring ER included “Tried to understand the other person’s concerns about COVID-19,” “Tried to understand how the other person felt about COVID-19,” and “Tried to help the other person(s) by listening to their concerns about SARS.” Higher total scores for the scales indicated a greater likelihood of using wishing and helping others to manage stress.

#### Demographic variables

Demographic variables described characteristics of the students and their programme of study. Age, gender, year of study, and clinical experience were variables collected for student characteristics. Programme characteristics included geographical location of the institution, academic level of the programme, and professions. Age was measured on a continuous scale, while the other variables were measured via categories.

All scales and demographic information were translated into Chinese for collecting the data in Putian. They were first translated into Chinese by a research team member and backward translated by a nurse educator. A dentist who is a native English speaker confirmed that the two versions of the scales contained the same meaning. The Chinese scales were then validated by five experts in nursing education, psychology, infection control, epidemiology, and public health nursing. The content validity of all scales ranged from very good to satisfactory (CVI = 0.7-1). Table 1 provides a summary of the scales included together with their psychometric properties. Psychometric analyses were conducted for each scale used in this study. First, confirmatory factor analysis was performed to assess the underlying structure of each scale. Internal consistency (Cronbach’s alpha) was calculated for each scale and its potential factors. In summary, our psychometric testing indicated that the study scales had good internal consistency, interpretability, and adequate factor structure. As a result, the scales were considered reliable, valid measures of the variables in the study.

### Statistical analysis

AMOS 23 was used to conduct analyses. For all tests, we used 2-sided p-values with alpha <=0.05 as the level of significance. We estimated the mean and standard deviation or frequency and percentage of the study variables. Missing values, normality, and outliers were first checked prior to the main analysis. No missing values nor outliers were found in the dataset. The dataset did not deviate from multivariate normality, as the Mardia value obtained was lower than the Mardia’s coefficient cutoff of 120 with 10 observed variables.^40^ For the main analysis, we ran a path analysis to explore the associations between covariates and outcomes. Maximum likelihood (ML) was used to estimate the regression coefficient of each path presented in the model. The model was trimmed until all insignificant paths were eliminated. Path coefficients were computed, including standardised and unstandardised regression coefficients with a 95% confidence interval. The model fit was determined by fit indices that included goodness-of-fit index (GFI) > 0.9,^41^ comparative fit index (CFI) and Tucker-Lewis index (TLI) >=0.9,^42,43^ root mean square error of approximation (RMSEA) < 0.05, and relative chi-square (X^2^/df) < 5.^44^

Subgroup analyses were conducted for geographical location, gender, professions, and clinical experience, as the former two demographic variables were reported to have a significant impact on health behaviours, while the latter two might affect a person’s knowledge about the infection.^10,12-15^ A chi-square test was performed to examine the difference of categorical socio-demographic variables between region, gender, professions, and clinical experience, while an independent t-test or one-way ANOVA test with Tukey post hoc test was employed for the continuous independent variables and the outcomes.

## Results

### Socio-demographic characteristics

Responses from a total of 2,706 students from healthcare professions were analysed. Most participants were from Fujian. The average age was 20.7 (SD=1.72). Out of the total number, 90.5% were female. About half were pursuing an associate degree, and 46.2% were studying for a bachelor’s degree, with around 90% studying nursing. About 70% of the participants were year 1 or year 2 students. More than 60% did not have clinical experience. In general, the PT scores of the students averaged 9.7 (SD=2.72), while their mean PS1 and mean PS2 scores were 12.3 (SD=4.68) and 7.2 (SD=2.72), respectively. Regarding the coping responses, the mean WT score was 6.8 (SD=1.16), and the mean ER score was 18.4 (SD=2.85). The mean SoD score was 43.4 (SD=8.18), and that for PHM was 23.1 (SD=2.86). The mean scores for PT, PS1, and PS2 were around the midpoint of the corresponding score ranges, indicating that the participants felt moderate threat and stress regarding COVID-19 and had moderate confidence about their ability to manage the situation. The mean scores for both wishful thinking and empathetic responding were on the high side of the corresponding score ranges, indicating that the respondents often used wishful thinking and empathetic responding to manage their stress. The participants revealed high compliance with SoD and PHM, as the high mean SoD and PHM scores reflected. Table 2 outlines the descriptive findings of the study.

**Table 2.** Sample characteristics and study variables by geographical location

|  | Hong Kong | Fujian | Total |  |
| --- | --- | --- | --- | --- |
|  | (N=462) | (N=2244) | (N=2706) |  |
| | n(%) / M±SD | n(%) / M±SD | n(%) / M±SD | p-value |
| Age | 21.1 ± 2.79 | 20.6 ± 1.38 | 20.7 ± 1.72 | <0.001*** |
| Gender |  |  |  | 0.032* |
| Female | 344(74.5) | 2105(93.8) | 2449(90.5) |  |
| Male | 118(25.5) | 139(6.2) | 257(9.5) |  |
| Academic level of the studying programme |  |  |  | <0.001*** |
| Diploma | 1(0.2) | 0(0.0) | 1(0.0) |  |
| Higher diploma | 80(17.3) | 0(0.0) | 80(3.0) |  |
| Associate degree | 2(0.4) | 1372(61.1) | 1374(50.8) |  |
| Bachelor degree | 379(82.0) | 872(38.9) | 1251(46.2) |  |
| Professions |  |  |  | <0.001*** |
| Nursing | 397(85.9) | 2099(93.5) | 2496(92.2) |  |
| Medical Laboratory Science | 22(4.8) | 48(2.1) | 70(2.6) |  |
| Chinese Medicine | 0(0.0) | 48(2.1) | 48(1.8) |  |
| Radiation Therapy | 11(2.4) | 21(0.9) | 32(1.2) |  |
| Pharmacy | 0(0.0) | 28(1.2) | 28(1.0) |  |
| Occupational Therapy | 18(3.9) | 0(0.0) | 18(0.7) |  |
| Physiotherapy | 14(3.0) | 0(0.0) | 14(0.5) |  |
| Year of study |  |  |  | <0.001*** |
| Year 1 | 147(31.9) | 963(42.9) | 1110(41.0) |  |
| Year 2 | 102(22.1) | 759(33.8) | 861(31.8) |  |
| Year 3 | 110(23.9) | 485(21.6) | 595(22.0) |  |
| Year 4 | 61(13.2) | 37(1.6) | 98(3.6) |  |
| Year 5 | 41(8.9) | 0(0.0) | 41(1.5) |  |
| Clinical experience |  |  |  | <0.001*** |
| No clinical experience | 189(40.9) | 1512(67.4) | 1701(62.9) |  |
| Less than 12 weeks | 127(27.5) | 254(11.3) | 381(14.1) |  |
| More than 12 weeks | 146(31.6) | 478(21.3) | 624(23.1) |  |
| *Study variables* |  |  |  |  |
| Perceived threat | 13.1 ± 2.59 | 9.0 ± 2.18 | 9.7 ± 2.72 | <0.001*** |
| Perceived stress |  |  |  |  |
| Domain 1 | 14.4 ± 4.64 | 11.9 ± 4.57 | 12.3 ± 4.68 | <0.001*** |
| Domain 2 | 7.0 ± 2.84 | 7.3 ± 2.75 | 7.2 ± 2.72 | 0.031* |
| Coping responses |  |  |  |  |
| Wishful thinking | 6.8 ± 1.40 | 6.9 ± 1.10 | 6.8 ± 1.16 | 0.348 |
| Empathetic responding | 17.9 ± 3.68 | 18.6 ± 2.64 | 18.4 ± 2.85 | <0.001*** |
| COVID-19 ICP behaviour |  |  |  |  |
| Social distancing | 41.6 ± 6.37 | 43.6 ± 8.47 | 43.3 ± 8.18 | <0.001*** |
| Personal hygiene measures | 22.6 ± 2.51 | 23.1 ± 2.92 | 23.1 ± 2.86 | <0.01** |

\* p<0.05; \*\* p<0.01; \*\*\* p<0.001

### Path analyses

The hypothesised model with all paths were tested first. Gradually, all non-significant paths were dropped until only significant paths remained. Figure 2 depicts the final predictive model with significant pathways and correlations. The trimmed path model shows good fit indexes, with chi-square *X*^2^=27.27, df=17, P=0.044; *X*^2^/df=1.61; GFI=0.998, CFI=0.997, TLI=0.992, RMSEA=0.015.

**Figure 2.**
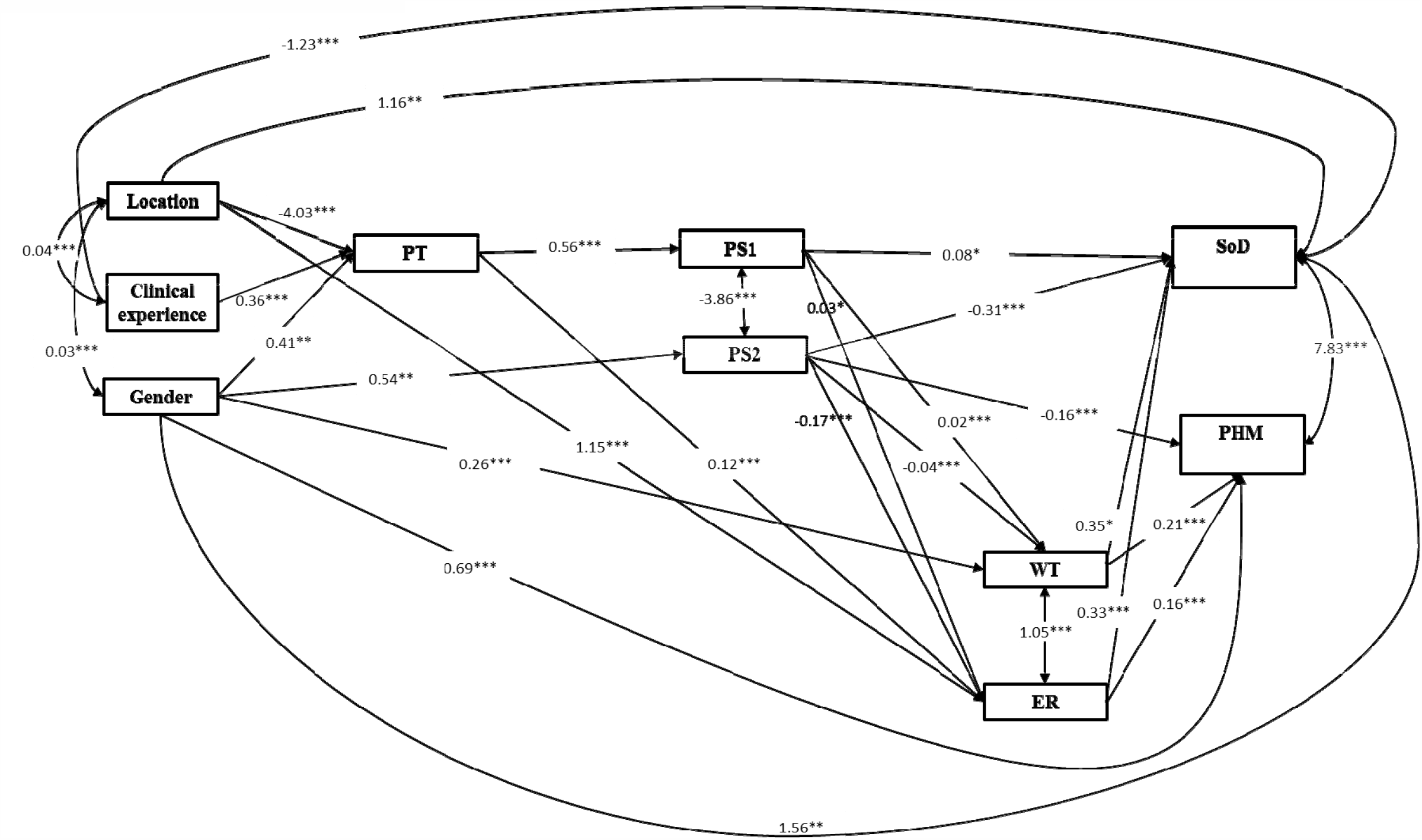
The final predictive model Location=Geographical location; PT=Perceived threat; PS1=Perceived stress domain 1; PS2=Perceived stress domain 2; WT=Wishful thinking; ER=Empathetic responding; SoD=Social distancing; PHM=Personal hygiene measures. Single arrow=unstandardized regression coefficient; Doubled arrow=correlation coefficient * p<0.05; ** p<0.01; *** p<0.001

Table 3 presents path coefficients with their standard errors and their significant levels for the model. The variables, which had significant direct and/or indirect effects on SoD and PHM, included gender, geographical location, clinical experience, perceived threat, perceived stress, and coping responses. PS1, PS2, WT, and ER were significantly associated with SoD. Among them, PS2, WT, and ER displayed the greatest contributions to SoD, with unstandardised regression coefficients (*β*) of −0.31 (95%CI=-0.37 to −0.19, p<0.001), 0.35 (95%CI=0.20 to 0.62, p=0.015), and 0.33 (95%CI=0.27 to 0.44, p<0.001), respectively. Fujian students had a higher mean SoD score than those in Hong Kong by 1.16 scores (95%CI=0.73 to 1.97). This result indicates that compared to Hong Kong students, Fujian students were more willing to engage in social distancing. Female students showed higher compliance with SoD than their male counterparts, with β of 1.56 (95%CI=0.99 to 2.62, p=0.004). Those having clinical experience had a lower mean SoD score than those without (β =-1.23, 95%CI=-1.83 to −0.91, p<0.001). These results imply that students with clinical experience are less likely to avoid going to public places than students without such experience.

**Table 3.** Regression coefficients of the significant paths in the final predictive model^#^

| Paths | | | B | $\beta$ | SE | 95% CI | p-value |
| --- | --- | --- | --- | --- | --- | --- | --- |
| Gender | → | PT | 0.044 | 0.405 | 0.152 | [ 0.244 , 0.703 ] | 0.008 |
| Location | → | PT | -0.558 | -4.029 | 0.121 | [ -4.157 , -3.792 ] | <0.001 |
| Experience | → | PT | 0.064 | 0.358 | 0.091 | [ 0.180 , 0.454 ] | <0.001 |
| PT | → | PS1 | 0.326 | 0.559 | 0.03 | [ 0.527 , 0.618 ] | <0.001 |
| Gender | → | PS2 | 0.057 | 0.537 | 0.172 | [ 0.355 , 0.874 ] | 0.002 |
| Gender | → | WT | 0.067 | 0.264 | 0.071 | [ 0.189 , 0.403 ] | <0.001 |
| PS1 | → | WT | 0.095 | 0.023 | 0.005 | [ 0.018 , 0.033 ] | <0.001 |
| PS2 | → | WT | -0.088 | -0.037 | 0.008 | [ -0.045 , -0.021 ] | <0.001 |
| Location | → | ER | 0.151 | 1.146 | 0.162 | [ 0.974 , 1.464 ] | <0.001 |
| PT | → | ER | 0.111 | 0.117 | 0.023 | [ 0.093 , 0.162 ] | <0.001 |
| PS2 | → | ER | -0.162 | -0.166 | 0.02 | [ -0.187 , -0.127 ] | <0.001 |
| PS1 | → | ER | 0.044 | 0.027 | 0.013 | [ 0.013 , 0.052 ] | 0.033 |
| Gender | → | SoD | 0.056 | 1.559 | 0.54 | [ 0.987 , 2.617 ] | 0.004 |
| Location | → | SoD | 0.053 | 1.164 | 0.41 | [ 0.729 , 1.968 ] | 0.005 |
| Experience | → | SoD | -0.073 | -1.233 | 0.303 | [ -1.827 , -0.912 ] | <0.001 |
| PS1 | → | SoD | 0.044 | 0.078 | 0.033 | [ 0.043 , 0.143 ] | 0.018 |
| PS2 | → | SoD | -0.104 | -0.308 | 0.059 | [ -0.371 , -0.192 ] | <0.001 |
| WT | → | SoD | 0.049 | 0.346 | 0.142 | [ 0.195 , 0.624 ] | 0.015 |
| ER | → | SoD | 0.113 | 0.326 | 0.058 | [ 0.265 , 0.440 ] | <0.001 |
| Gender | → | PHM | 0.071 | 0.69 | 0.18 | [ 0.499 , 1.043 ] | <0.001 |
| PS2 | → | PHM | -0.151 | -0.156 | 0.019 | [ -0.176 , -0.119 ] | <0.001 |
| WT | → | PHM | 0.086 | 0.212 | 0.049 | [ 0.160 , 0.308 ] | <0.001 |
| ER | → | PHM | 0.161 | 0.161 | 0.02 | [ 0.140 , 0.200 ] | <0.001 |

<sup>#</sup>Model fit indices: $\chi^2 = 27.270$ ; $df = 17$ ; $p = 0.044$ ; $\chi^2/df = 1.604$ ; GFI = 0.998; CFI = 0.997; TLI = 0.992; RMSEA = 0.015
Experience=Clinical experience; Location=Geographical location; PT=Perceived threat; PS1=Perceived stress domain 1; PS2=Perceived stress domain 2; WT=Wishful thinking; ER=Empathetic responding; SoD=Social distancing; PHM=Personal hygiene measures
B=Standardized regression estimates; $\beta$ =Unstandardized regression estimates; SE=Standard error; CI=Confidence interval

Similar to SoD, PS2 (β=-0.156, 95%CI=-0.18 to −0.12, p<0.001), WT (*β*=0.21, 95%CI=0.16 to 0.31, p<0.001), and ER (β=0.16, 95%CI=0.14 to 0.2, p<0.001) were the three variables with the greatest contributions towards PHM, though they exhibited lesser effects compared to those on SoD. PS1 exerted an indirect impact on PHM through WT and ER. Gender was the only significant demographic variable that had a direct impact on PHM (*β*=0.69, 95%CI=0.50 to 1.04, p<0.001), in that female students were more likely to comply with personal hygiene measures than male students. The negative associations of PS2 with SoD and PHM indicate that a high level of confidence about managing the current situation increased compliance with SoD and PHM. Unlike the hypothesised model, PT did not have a direct impact on both SoD and PHM but infiltrated its impact into the two outcomes by way of PS1 and ER.

The three demographic variables and PS2 exhibited their indirect influences on SoD and PHM through other variables in addition to their direct effects. Geographical location was positively associated with PT and ER, with *β* of -4.03 (95%CI=-4.16 to -3.79, p<0.001) and 1.15 (95%CI=0.97 to 1.46, p< 0.001), respectively. This result means that Fujian students perceived less threat and adopted more empathetic responding than Hong Kong students. Gender positively associated with PT (*β*=0.41, 95%CI=0.24 to 0.70, p=0.008), PS2 (*β*=0.54, 95%CI=0.36 to 0.87, p=0.002), and WT (*β*=0.26, 95%CI=0.19 to 0.40, p<0.001). Students having clinical experience had a higher PT score than those without (*β*=-1.23, 95%CI=-1.83 to −0.91, p<0.001), indicating that students with clinical experience tended to feel more threat than those without. Students with higher confidence towards managing the current situation drew on wishful thinking and empathetic responding to cope with the situation as reflected in the significant negative association between PS2 and WT *(β*=-0.04, 95%CI=-0.05 to −0.02, p<0.001), and between PS2 and ER (*β*=-0.17, 95%CI=-0.19 to −0.13, p<0.001).

### Subgroup analyses

The Hong Kong students exhibited significantly higher PT (p<0.001) and PS1 scores (p<0.001) but a lower PS2 score (p<0.05) than their Fujian counterparts. The Fujian students had a significantly higher ER score and were more likely to comply with SoD and PHM than the Hong Kong students (Table 2). Female students had a lower PT score and higher PS2, WT, SoD, and PHM scores in comparison to male students (Table 4). Students with no clinical experience revealed significantly lower PT and PS1 scores than those with clinical experience (p<0.001). Participants without clinical experience were found to practise higher compliance with SoD compared to the other two groups (p<0.001) (Table 5). Physiotherapy students had the highest PT score, followed by occupational therapy students. The same pattern was observed for PS1, though it is not statistically significant (Table 6). In summary, Fujian female students, irrespective of profession, showed greater compliance with SoD and PHM. Students with no clinical experience adhered more to SoD compared to those having clinical experience.

**Table 4.** Study variables by gender

|  | Male | Female | Total |  |
| --- | --- | --- | --- | --- |
|  | (N=257) | (N=2499) | (N=2706) |  |
|  | M±SD | M±SD | M±SD | p-value |
| Perceived threat | 10.5 ± 3.50 | 9.6 ± 2.61 | 9.7 ± 2.72 | <0.001*** |
| Perceived stress |  |  |  |  |
| Domain 1 | 12.6 ± 5.39 | 12.3 ± 4.60 | 12.3 ± 4.68 | 0.485 |
| Domain 2 | 6.8 ± 3.21 | 7.3 ± 2.71 | 7.2 ± 2.77 | 0.019* |
| Coping responses |  |  |  |  |
| Wishful thinking | 6.6 ± 1.45 | 6.9 ± 1.12 | 6.8 ± 1.16 | 0.013* |
| Empathetic responding | 18.3 ± 3.68 | 18.5 ± 2.75 | 18.4 ± 2.85 | 0.598 |
| COVID-19 ICP behaviour |  |  |  |  |
| Social distancing | 41.6 ± 8.72 | 43.5 ± 8.11 | 43.3 ± 8.18 | 0.001** |
| Personal hygiene measures | 22.4 ± 3.64 | 23.1 ± 2.76 | 23.1 ± 2.86 | 0.004** |

\* p<0.05; \*\* p<0.01; \*\*\* p<0.001

**Table 5.** Study variables by clinical experience

|  | No Clinical<br>Experience | Less than 12<br>weeks | More than 12<br>weeks | Total |  |
| --- | --- | --- | --- | --- | --- |
|  | (N=1701) | (N=381) | (N=624) | (N=2706) |  |
|  | M±SD | M±SD | M±SD | M±SD | p-value |
| Perceived threat | 9.3 ± 2.59 | 10.6 ± 2.87 | 10.2 ± 2.76 | 9.7 ± 2.72 | <0.001*** |
| Perceived stress |  |  |  |  |  |
| Domain 1 | 12.1 ± 4.61 | 12.8 ± 4.94 | 12.8 ± 4.65 | 12.3 ± 4.68 | 0.001** |
| Domain 2 | 7.3 ± 2.7 | 7.0 ± 2.88 | 7.2 ± 2.87 | 7.2 ± 2.77 | 0.084 |
| Coping responses |  |  |  |  |  |
| Wishful thinking | 6.8 ± 1.14 | 6.8 ± 1.21 | 6.8 ± 1.16 | 6.8 ± 1.16 | 0.928 |
| Empathetic responding | 18.5 ± 2.77 | 18.4 ± 2.99 | 18.4 ± 2.98 | 18.4 ± 2.85 | 0.874 |
| COVID-19 ICP behaviour |  |  |  |  |  |
| Social distancing | 43.7 ± 7.91 | 42.6 ± 8.68 | 42.5 ± 8.53 | 43.3 ± 8.18 | 0.001** |
| Personal hygiene measures | 23.0 ± 2.92 | 23.1 ± 2.69 | 23.1 ± 2.81 | 23.1 ± 2.86 | 0.448 |

\*\* p<0.01; \*\*\* p<0.001

**Table 6.** Study variables by professions

|  | **Nursing** | **Medical Laboratory Science** | **Chinese Medicine** | **Radiation Therapy** | **Pharmacy** | **Occupational Therapy** | **Physiotherapy** | **Total** |  |
| --- | --- | --- | --- | --- | --- | --- | --- | --- | --- |
|  | **(N=2496)** | **(N=70)** | **(N=48)** | **(N=32)** | **(N=28)** | **(N=18)** | **(N=14)** | **(N=2706)** |  |
|  | **M±SD** | **M±SD** | **M±SD** | **M±SD** | **M±SD** | **M±SD** | **M±SD** | **M±SD** | **p-value** |
| Perceived threat | 9.7 ± 2.69 | 10 ± 3.3 | 8.3 ± 2.04 | 10.3 ± 2.82 | 8.5 ± 2.15 | 11.8 ± 3.25 | 13.3 ± 1.94 | 9.7 ± 2.72 | <0.001*** |
| Perceived stress |  |  |  |  |  |  |  |  |  |
| Domain 1 | 12.4 ± 4.66 | 12.6 ± 4.33 | 11.4 ± 5.26 | 11.9 ± 5.48 | 10.6 ± 4.72 | 13.5 ± 4.41 | 14.9 ± 4.51 | 12.3 ± 4.68 | 0.063 |
| Domain 2 | 7.2 ± 2.76 | 7.1 ± 2.85 | 7.4 ± 3.26 | 7.1 ± 2.71 | 7.4 ± 2.64 | 7.2 ± 2.41 | 7.1 ± 2.34 | 7.2 ± 2.77 | 0.998 |
| Coping responses |  |  |  |  |  |  |  |  |  |
| Wishful thinking | 6.8 ± 1.16 | 7.1 ± 1.07 | 7.1 ± 1.25 | 6.9 ± 1.13 | 6.6 ± 1.06 | 6.4 ± 1.34 | 6.6 ± 1.55 | 6.8 ± 1.16 | 0.196 |
| Empathetic responding | 18.4 ± 2.85 | 19.1 ± 3.04 | 18.4 ± 2.89 | 18.3 ± 2.73 | 18.2 ± 3.02 | 18 ± 3.05 | 17.7 ± 1.27 | 18.4 ± 2.85 | 0.495 |
| COVID-19 ICP behaviour |  |  |  |  |  |  |  |  |  |
| Social distancing | 43.3 ± 8.27 | 44.3 ± 5.06 | 42.9 ± 9.48 | 43.1 ± 6.11 | 43.0 ± 7.93 | 39.6 ± 8.02 | 42.5 ± 4.74 | 43.3 ± 8.18 | 0.555 |
| Personal hygiene measures | 23.1 ± 2.89 | 23.3 ± 1.94 | 23.3 ± 2.93 | 22.1 ± 2.85 | 22.8 ± 2.99 | 22.1 ± 1.78 | 23.7 ± 1.33 | 23.1 ± 2.86 | 0.284 |

\*\*\* p<0.001

## Discussion

In this study, the use of path analysis to construct the predictive model revealed the direct and indirect contributions of demographic and independent variables to compliance with social distancing and personal hygiene measures. The final predictive model for COVID-19 ICPs demonstrates that the significant demographic variables (i.e. gender, geographical location, clinical experience) and stress level do not merely contribute to social distancing and personal hygiene measures directly but also impose indirect effects on them through other studied variables in the model. Perceived threat, as expected, does not have a direct effect on compliance with COVID-19 ICPs. Wishful thinking and empathetic responding directly and positively contribute to the two outcomes.

### Demographic characteristics

The significant associations of gender and geographical location with infection control measures reported in this study were similar to those in previous studies in which females showed greater compliance with infection control measures such as handwashing than males among college students and healthcare workers.^20,33-35^ Healthcare workers in different countries were reported to have varying adherence to personal protection measures.^36-38^ The contributions of the present study are twofold. First, the negative association between clinical experience and compliance with social distancing suggests that healthcare students with clinical experience comply less with social distancing. Second, geographical location and clinical experience directly and positively associate with compliance with social distancing only, though not personal hygiene measures. The higher compliance with social distancing found in Fujian students might be the result of a difference in regulations regarding social distancing between Hong Kong and Fujian. Restrictive measures were launched at an early stage of the COVID-19 outbreak in Fujian. As a consequence, the Fujian government imposed the strictest measures to control social contact among people, such as banning social gatherings and suspending public transportation. In contrast to Fujian, the regulations affecting social distancing in Hong Kong have been less restrictive at that time. For instance, in Hong Kong, people could have gatherings, though these were limited to four attendees at most. Public transport was still in operation, but passengers were required to put on facemasks. In addition, students and workers in Mainland China are used to travelling across cities and provinces seeking better study and job opportunities. Residents of Fujian may thus be more vigilant about the potential for infection caused by contact in public areas with strangers from high-risk cities. Regarding the negative association of clinical experience and compliance with social distancing, we should interpret it with caution, as students with clinical experience were mainly clustered in the Hong Kong samples. As Hong Kong students were found to have significantly lower compliance with social distancing than the Fujian students in the subgroup analysis, the negative association of clinical experience with compliance in social distancing might be biased by the skewed samples. Further studies shall be launched to confirm this association.

### Perceived stress

PSS-10 is a widely used scale to assess the perception of stress in response to life events in an adult population. Unlike previous studies that commonly used the total scale score for analysis, we split the scale into two domains – stress level (PS1) and confidence about managing the present situation (PS2) – when performing the path analysis. The present model suggests that confidence about managing the situation has a stronger predictive effect than one’s stress level for both social distancing and personal hygiene measures. Confidence about managing an encountered difficulty relates to one’s sense of self-efficacy.^39^ This sense is a major behaviour-specific cognitive factor contributing to engagement in behaviour that promotes health. A high level of self-efficacy can reduce the perception of barriers, improving the likelihood of engaging in healthful practices.^40^ Public health interventions may focus on enhancing people’s self-efficacy to increase adherence to ICPs for COVID-19.

### Coping responses

Wishful thinking and empathetic responding were both found to positively associate with social distancing and personal hygiene measures, results that are incongruent with those from previous studies regarding SARS and other epidemics.^25,26^ In this study, wishful thinking and empathetic responding both directly and positively contributed to adherence to social distancing and personal hygiene measures. Conversely, Lee-Baggley et al.^25^ reported that wishful thinking did not relate to personal hygiene measures in SARS samples. The distinctive nature of COVID-19 may explain the incongruent findings between previous studies and ours. The transmissibility of COVID-19 is much higher than that for viruses encountered in previous incidents. The progress of the current virus is fluctuating, and no medications are available at this time to cure the infection. People are in fact facing many unknowns and uncertainties that exert a significant impact on their psychological wellbeing. Recent studies have reported that COVID-19 has compromised the psychological wellbeing of healthcare workers and the general public.^41-43^ People suffering from despair may turn to wishful thinking to relieve their psychological burdens in an attempt to preserve their mental health. Although the literature often associates wishful thinking with higher stress level and poorer health outcomes,^27,44,45^ the use of wishful thinking in the face of the COVID-19 pandemic may have a protective effect on people’s mental wellbeing.^46^ The positive correlation between wishful thinking and empathetic responding may support the protective effect of wishful thinking on the participants’ mental health in that greater employment of wishful thinking would simultaneously increase the use of a proactive stress-coping strategy – empathetic responding to manage the stress. In addition, the participants with more confidence about managing the threat practise more wishful thinking as reflected by the negative association between PS2 and this type of mental focus.

The present study showed that empathetic responding had a greater influence on social distancing than on personal hygiene measures, as the unstandardised regression coefficients of the corresponding paths in the model reflected. Conversely, previous studies found that empathetic responding had a higher association with personal hygiene measures in SARS and West Nile virus samples.^29,30^ The different findings among studies may be explained by the differences in the key strategies to control viral spread in different outbreaks. In COVID-19, social distancing is a key strategy to mitigate community spread of the infection, along with personal hygiene measures. However, in the case of SARS, personal hygiene measures were emphasised to quell the outbreak. Empathetic responding thereby relates to key specific public health strategies to control the corresponding outbreaks. Another possible explanation for the incongruence may involve the sample characteristics in the present study. In our samples, over 80 percent of the participants were from Fujian. This number is significant because the Fujian group was found to have a significantly higher mean empathetic responding score than the Hong Kong group in subgroup analysis. The greater use of empathetic responding observed in the Fujian group may be attributable to the social norm to maintain close relationships within neighbourhoods and support each other in Mainland China. Although public health measures to control COVID-19 have forced the participants to isolate themselves from others with whom they used to be in close contact, they may continue to interact as usual through other means such as communication apps or social media to share their stress and difficulties. Further, the Mainland students are accustomed to leaving their hometowns to seek better study opportunities in more prosperous cities. Because some of the students were forced to stay in hostels during the outbreak because of the lockdown and suspension of transportation, they may exhibit empathetic behaviours to support each other to transcend a difficult situation.

### Strengths, Limitations and Recommendations

This study is the first to use path analysis to explore factors associated with compliance with infection control measures for the control of COVID-19 among healthcare students. Both direct and indirect effects of potential factors contributing to compliance with COVID-19 ICPs were revealed. This approach is more advantageous than regression analysis in the sense that any indirect effects of the factors would not be masked in path analysis. The statistically significant paths and the good fit of the model provide empirical support for the study hypotheses. Stakeholders may refer to the present model to provide interventions targeted not only at factors directly associated with social distancing and personal hygiene measures but also factors associated with confidence in managing the situation and coping responses that can improve compliance.

Although this study contributes to the knowledge of factors associated with compliance with COVID-19 ICPs, several limitations may limit the validity of the findings. First, the convenience and snowball samples may not reflect the full characteristics of the entire population. For instance, the participants were mainly from junior years, and only a few senior students who would have more clinical experience were included. Thus, the negative association found between clinical experience and compliance with social distancing may be biased because the samples were dominated by junior students. Second, since the data collection was conducted in China, the predictive model may not apply to other countries with different socio-cultural backgrounds and dissimilar implementation policies for COVID-19 ICPs. Third, a self-reported survey was used to collect students’ perceptions and likelihood to comply with COVID-19 ICPs. Thus, the data may be subject to social response bias. Fourth, the present model was established based on cross-sectional data, making it difficult to draw causal inferences about the relationships between variables. A world-wide study should be designed to collect subjects from more countries to enhance study validity. Also, the use of random sampling and inclusion of more objective data collection methods should be considered to minimise biases caused by unbalanced sample characteristics and subjective responses. Further research should additionally examine the causal relationships using longitudinal methodologies.

### Conclusion

The predictive model developed in this study is the first to explore factors associated with compliance with infection control measures by healthcare students amid the COVID-19 outbreak. The findings help stakeholders to identify students at risk for poor compliance. The findings suggest that students who are male, reside in Hong Kong, have more clinical experience, and exhibit weak confidence about managing the threat tend to display lower compliance in terms of social distancing and personal hygiene measures. Wishful thinking, which has been generally connected to negative health outcomes,^28^ was found to be a positive factor in complying with infection control measures for COVID-19. Public health interventions should target groups susceptible to poor compliance to strengthen the control of COVID-19 spread.

## Data Availability

The data file may be provided upon reasonable request.

## Notes

### Competing Interest Statement

The authors have declared no competing interest.

### Clinical Trial

it is a cross sectional study collecting data with online survey.

### Funding Statement

no funder/sponsor

### Author Declarations

The study protocol, questionnaires and procedures were approved by the corresponding institutional review boards in Hong Kong and Fujian: Tung Wah College (REC2020056) and Putian College (2020-42).

## References

1. World Health Organization. WHO Coronavirus Disease (COVID-19) Dashboard. June 30, 2020. https://covid19.who.int/

2. Special Expert Group for Control of the Epidemic of Novel Coronavirus Pneumonia of the Chinese Preventive Medicine. An update on the epidemiological characteristics of novel coronavirus pneumonia (COVID-19). Chinese Journal of Epidemiology. 2020;41(2):139–144.

3. Centers for Disease Control and Prevention. How COVID-19 spreads. June 16, 2020. https://www.cdc.gov/coronavirus/2019-ncov/prevent-getting-sick/how-covid-spreads.html?CDC_AA_refVal=https%3A%2F%2Fwww.cdc.gov%2Fcoronavirus%2F2019-ncov%2Fprepare%2Ftransmission.html

4. World Health Organization. Coronavirus disease (COVID-19) Advice for The Public. June 4, 2020. https://www.who.int/emergencies/diseases/novel-coronavirus-2019/advice-for-public

5. United Nation. COVID-19 likely to shrink global GDP by almost one percent in 2020. April 1, 2020. https://www.un.org/sustainabledevelopment/blog/2020/04/covid-19-likely-to-shrink-global-gdp-by-almost-one-per-cent-in-2020/

6. France-Presse A. Coronavirus kills 20.5 million US jobs in April in historic collapse. World/United State & Canada, South China Morning Post. May 8, 2020. https://www.scmp.com/news/world/united-states-canada/article/3083588/coronavirus-kills-205-million-us-jobs-april

7. BBC News. Huge rise in people claiming unemployment benefit. May 19, 2020. https://www.bbc.com/news/business-52719230

8. Wilder-Smith A, Chiew CJ, Lee VJ. Can we contain the COVID-19 outbreak with the same measures as for SARS? The Lancet 2020;20(5):e102-e107. doi:https://doi.org/10.1016/S1473-3099(20)30129-8

9. Centre for Health Protection, Department of Health HKSAR. Guidelines on Prevention of COVID-19 for the General Public. May 25, 2020 https://www.chp.gov.hk/files/pdf/nid_guideline_general_public_en.pdf

10. Bish A, Michie S. Demographic and attitudinal determinants of protective behaviours during a pandemic: A review. Br J Health Psychol. 2010;15(4):797-824. doi: https://doi.org/10.1348/135910710X485826

11. Marshall H, Tooher R, Collins J, et al. Awareness, anxiety, compliance: Community perceptions and response to the threat and reality of an influenza pandemic. Am J of Infec Control. 2012;40(3):270-272. doi: https://doi.org/10.1016/j.ajic.2011.03.015

12. Leung GM, Quah S, Ho LM, et al. A tale of two cities: Community psychobehavioral surveillance and related impact on outbreak control in Hong Kong and Singapore during the Severe Acute Respiratory Syndrome Epidemic. Infect Control & Hosp Epidemiol. 2004;25(12):1033-1041. doi: https://doi.org/10.1086/502340

13. Tooher R, Collins JE, Street JM, Braunack-Mayer A, Marshall H. Community knowledge, beahviours and attitudes about the 2009 H1N1 Influenza pandemic: A systematic review. Influenza Other Respir Viruses. 2013;7(6):1316-1327. doi: https://doi.org/10.1111/irv.12103

14. Eastwood K, Durrheim D, Francis JL, et al. Knowledge about pandemic influenza and compliance with containment measures among Australians. Bulletin of the World Health Organization. 2009;87(8):588–594. https://www.scielosp.org/pdf/bwho/2009.v87n8/588-594/en

15. Bults M, Beaujean DJMA, Richardus JH, Voeten HACM. Perceptions and behavioral responses of the general public during the 2009 influenza (H1N1) pandemic: A systematic review. Disaster Med Public Health Prep. 2015;9(2): 207-219. doi: https://doi.org/10.1017/dmp.2014.160

16. Powell-Jackson T, King JJC, Makungu, C, et al. Infection prevention and control compliance in Tanzanian outpatient facilities: A cross sectional study with implications for the control of COVID-19. The Lancet. 2020;8(6):e780-e789. doi: https://doi.org/10.1016/S2214-109X(20)30222-9

17. Sigayeva A, Green K, Raboud M, et al. Factors associated with critical-care healthcare workers’ adherence to recommended barrier precautions during the Toronto Severe Acute Respiratory Syndrome Outbreak. Infect Control Hosp Epidemiol. 2007;28(11): 1275-1283. doi: https://doi.org/10.1086/521661

18. Moore D, Gamage B, Bryce E, Copes R, Yassi A. Protecting health care workers from SARS and other respiratory pathogens: Organizational and individual factors that affect adherence to infection control guidelines. Am J Infect Control. 2005;33(2):88-96. doi: https://doi.org/10.1016/j.ajic.2004.11.003

19. Chor JSY, Pada SK, Stephenson I, et al. Differences in the compliance with hospital infection control practices during the 2009 influenza H1N1 pandemic in three countries. J Hosp Infect. 2012; 81(2): 98-103. doi:https://doi.org/10.1016/j.jhin.2012.04.003

20. Brooks SK, Greenberg N, Wessely S, Rubin GJ. Factors affecting healthcare workers’ compliance with social and behavioural infection control measures during emerging infectious disease outbreaks: Rapid evidence review. medRxiv. May 29, 2020. doi: https://doi.org/10.1101/2020.05.27.20114744

21. Pittet D, Simon A, Hugonnet S, Pessoa-Silva CL, Sauvan V, Pernger TV. Hand hygiene among physicians: Performance, beliefs, perceptions. Ann Intern Med 2004; 141: 1-8. doi: https://doi.org/10.7326/0003-4819-141-1-200407060-00008

22. Darawad MW, Al-Hussami M. Jordanian nursing students’ knowledge of, attitudes towards and compliance with infection control precautions. Nurse Educ Today.2013; 33(6): 580-583. doi: https://doi.org/10.1016/j.nedt.2012.06.009

23. Janz NK, Becker MH. The Health Belief Model: A Decade Later. Health Educ Q. 1984;11(1): 1–47. Doi: :10.1177/109019818401100101

24. Folkman S. Stress: appraisal and coping. In: Gellman MD, Turner JR, editors. Encyclopaedia of Behavioral Medicine. New York: Springer; 2013

25. Lee-Baggley D, DeLongis A, Voorhoeave P. Coping with the threat of severe acute respiratory syndrome: Role of threat appraisals and coping responses in health behaviors. Asian J Social Psychology. 2004; 7:9–23. doi: 10.1111/j.1467-839X.2004.00131.x

26. Putermna E, DeLongis A, Lee-Baggley D, Greenglass E. Coping and health behaviors in times of global health crises: Lessons from SARS and West Nile. Glo Public Health. 2009; 4(1): 69–81. doi: 10.1080/17441690802063304

27. Penley JA, Tomaka J, Wiebe JS. The association of coping to physical and psychological health outcomes: A meta-analytic review. J Behavioral Medicine. 2002; 25:551–603. Doi: https://doi.org/10.1023/A:1020641400589

28. Vandenbroucke JP, von Elm E, Altman DG, et al. Strengthening the Reporting of Observational Studies in Epidemiology [STROKE]: Explanation and Elaboration. PLoS Med. 2007;4:e297. Doi: https://doi.org/10.1371/journal.pmed.0040297

29. Kline RB. Principles and practice of structural equation modeling. 3rd ed. New York: The Guilford Press;2011.

30. Cohen S, Williamson G. Perceived stress in a probability sample of the United States In: Spacapam S, Oskamp S, editors. The social psychology of health. Newbury Park,CA: Sage; 1988. p. 31–67. 1988

31. Folkman S, Lazarus RS. The relationship between coping and emotion: implications for theory and research. Soc Sci Med. 1988;26(3):309–317. Doi: https://doi.org/10.1016/0277-9536(88)90395-4

32. O’Brien TB, DeLongis A. The interactional context of problem-, emotion-, and relationship-focused coping: The role of the big five personality factors. J Pers. 1996;64:775-813. doi: https://doi.org/10.1111/j.1467-6494.1996.tb00944.x

33. Anderson JL, Warren CA, Perez E, et al. Gender and ethnic differences in hand hygiene practices among college students. Am J Infect Control. 2008;36:5:361-368. doi: https://doi.org/10.1016/j.ajic.2007.09.007

34. White C, Kolble R, Carlson R, Lipson N. The impact of a health campaign on hand hygiene and upper respiratory illness among college students living in residence halls. J Am Coll Health. 2005;53(4):175–181. Doi: 10.3200/JACH.53.4.175-181

35. Powell-Jackson T, King JJC, Makungu C. Infection prevention and control compliance in Tanzanian outpatient facilities: A cross-sectional study with implications for the control for the control of COVID-19. The Lancet Global Health, 2020;8(6):e780-e789. doi: https://doi.org/10.1016/S2214-109X(20)30222-9

36. Chor JSY, Pada SK, Stephenson I, et al. Differences in the compliance with hospital infection control practices during the 2009 influenza H1N1 pandemic in three countries. J Hosp Infect. 2012;81(2):98-103. Doi: https://doi.org/10.1016/j.jhin.2012.04.003

37. Koh GC-H, Abikusno N, Kwing CS, et al. Avian influenza and South Jakarta primary healthcare workers: a controlled mixed-method study. Trop Med Int Health. 2009;14(7):817–29. Doi: :10.1111/j.1365-3156.2009.02297.x

38. Wong SYS, Wong W, Jaakkimainen L, Bondy S, Tsang KK, Lee A. Primary care physicians in Hong Kong and Canada – how did their practices differ during the SARS epidemic? Fam Pract. 2005;22(4):361-6. Doi: https://doi.org/10.1093/fampra/cmi036

39. Bandura A. Self-efficacy mechanism in human agency. Am Psychol. 1982;37(2):122–147. doi:10.1037/0003-066X.37.2.122.

40. Janz NK, Becker MH. The health belief model: A decade later. Health Educ Q. 1984;147(1984):1–7. Doi: 10.1177/109019818401100101

41. Lai J, Ma S, Wang Y, et al. Factors associated with mental health outcomes among health care workers exposed to coronavirus disease 2019. JAMA Network Open, 2020;3(3):e203976. doi: 10.1001/jamanetworkopen.2020.3976

42. Ma Y, Rosenheck R, He H. Psychological stress among health care professionals during the 2019 novel coronavirus disease outbreak: Cases from online consulting customers. Intensive and Critical Care Nursing. June 28, 2020. doi: https://doi.org/10.1016/j.iccn.2020.102905

43. Kuang J, Ashraf S, Das U, Bicchieri C. Awareness, risk perception, and stress during the COVID-19 pandemic in communities of Tamil Nadu, India. PsyArXiv. June 27, 2020. doi:10.31234/osf.io/qhgrd

44. Onieva-Zafra MD, Fernandez-Munoz JJ, Fernandex-Martinex E, et al. Anxiety, perceived stress and coping strategies in nursing students: A cross-sectional correlational descriptive study. Europe PMC. June 25, 2020. doi: 10.21203/rs.2.10722/v2

45. Quynh HHN, Tanasugarn C, Kengganpanich M, Lapvongwatana P, Long KQ, Truc TT. Mental Well-being, and Coping Strategies during Stress for Preclinical Medical Students in Vietnam. JPSS. 2020;2(2):116–129. doi: 10.25133/JPSSv28n2.008

46. Karaca, A., Yildirim, N., Cangur, S., Acikgoz, F., Akkus, D. Relationship between mental health of nursing students and coping, self-esteem and social support. Nurs Educ Today. 2019;76:44–50. Doi: https://doi.org/10.1016/J.NEDT.2019.01.029

